# Experience in the adoption of Research Electronic Data Capture (REDCap) as a tool for medical research in Tanzania

**DOI:** 10.1101/2025.10.25.25338769

**Authors:** Raphael Zozimus Sangeda, Upendo Masamu, Daniel Kandonga, Fredrick Mbuya, Liberata Mwita, Frank Makundi, Josephine Mgaya, Elisha Osati, Agnes Jonathan, Bruno P. Mmbando, Siana Nkya, Emmanuel Balandya, Julie Makani

**Affiliations:** Department of Pharmaceutical Microbiology, School of Pharmacy, Muhimbili University of Health and Allied Sciences, Dar es Salaam, Tanzania; Muhimbili Sickle Cell Program, Department of Hematology and Blood Transfusion, Muhimbili University of Health and Allied Sciences, Dar es Salaam, Tanzania; Muhimbili National Hospital, Dar es Salaam, Tanzania; National Institute for Medical Research, Tanga Center, Tanzania; Department of Biochemistry, Muhimbili University of Health and Allied Sciences (MUHAS), Dar-es-Salaam, Tanzania; Department of Physiology, Muhimbili University of Health and Allied Sciences, Dar es Salaam, Tanzania; Department of Hematology and Blood Transfusion, Muhimbili University of Health and Allied Sciences (MUHAS), Dar-es-Salaam, Tanzania

**Keywords:** Research Electronic Data Capture (REDCap), Data Collection, Data Management, Biomedical Research, Health Informatics, Capacity Building, Tanzania, Quality Control, Systems Integration

## Abstract

**Objective:** To document the deployment and training of Research Electronic Data Capture (REDCap) at Muhimbili University of Health and Allied Sciences (MUHAS) in Tanzania and to evaluate user satisfaction and perceived impact on research practices in Tanzania.

**Materials and Methods:** Six structured REDCap training workshops were conducted between 2016 and 2022. The post-training evaluation surveys were completed by 110 participants. Descriptive statistics were generated for demographics and cadres, while Likert-scale responses assessed satisfaction and perceived utility. Diverging stacked bar charts were used to visualize attitudes toward adopting REDCap.

**Results:** Most trainees were male (62.7%), aged 26–32 years (52.7%), and affiliated with MUHAS (66.4%), with additional representation from Ghana and Nigeria. The participants included academic or research staff (41.8%), postgraduate students (28.2%), undergraduate students (11.8%), and other professionals (18.2%). Post-training evaluations indicated consistently high satisfaction, with mean scores above 4.4 on a 5-point scale. Trainees strongly endorsed REDCap’s ability to enhance research effectiveness (mean 4.53, SD 0.65), increase productivity (mean 4.52, SD 0.69), and improve task completion (mean 4.50, SD 0.68). More than 85% of the respondents agreed or strongly agreed with the positive statements, underscoring the broad acceptance of REDCap as a reliable tool for research data management.

**Discussion:** The findings underscore the importance of structured training in mitigating barriers to adoption and enhancing data quality, efficiency, and institutional visibility.

**Conclusion:** Structured REDCap training in Tanzania was associated with high user satisfaction and perceived improvements in research productivity and data quality, offering a scalable model for strengthening data management capacity in sub-Saharan Africa.

## Introduction

Data interoperability and portability are critical for the design of health research projects. The ability to merge datasets from diverse sources depends on precision and validity, underscoring the importance of robust data quality management. This is particularly important in multi-country studies, where standardized approaches enable harmonized information exchange across sites [1–3]. Research Electronic Data Capture (REDCap) is a secure web-based application, free to non-profit institutions under license, that supports the creation and management of surveys and databases [4]. As of 2023, REDCap was used by 7,123 institutions in 156 countries, helping over two million projects and cited in more than 22,700 publications [4–6]. Its features include audit trails, double data entry for integrity checks, offline mobile data capture, and a randomization module to support unbiased allocation. Specialized services, such as the Data Transfer Service (DTS) and Twilio Telephony Services (TTSS), further expand functionality, enabling data exchange with electronic medical records and sending SMS-based survey invitations [5,7–9]. These advanced capabilities make REDCap well-suited for collaborative biomedical research, although they also require additional training and reliable Internet connectivity.

Compared with commonly used tools in Tanzania, such as Excel, SPSS, KoboToolbox, Google Forms, OpenClinica [10], and custom-built applications, REDCap offers distinct advantages in research security, project management, and standardization. Excel and Google Forms offer user-friendly entry but lack advanced safeguards. SPSS is powerful for analysis, but it is not designed for data collection workflows. KoboToolbox and Google Forms work well for basic surveys, but fall short for complex projects that require strict security. OpenClinica offers comparable trial management functionality but differs in usability and customization. Custom applications can be tailored; however, they are costly to develop and maintain. In contrast, REDCap integrates these capacities into a versatile, secure, and research-oriented platform [7–9].

Its adoption in Africa demonstrates the feasibility and impact of REDCap in resource-constrained environments. It has supported healthcare research, training, and infrastructure development [10,11]. However, data management inconsistencies persist in Tanzania. Ad hoc reliance on spreadsheets and paper forms has led to variable data quality, complicating data aggregation across studies and limiting broader research impact[12,13]. These challenges mirror broader gaps in data science capacity across Africa, where underinvestment in infrastructure and workforce development continues to hinder sustainable adoption of digital research tools [14].

This study documents the structured deployment of REDCap and training at the Muhimbili University of Health and Allied Sciences (MUHAS) in Tanzania, which was implemented in collaboration with regional and international partners. By reporting participant demographics, user satisfaction, and perceived impacts on efficiency and data quality, we provide empirical evidence on REDCap adoption in Tanzania, offering one of the first structured evaluations of training outcomes in sub-Saharan Africa.

### Significance Statement

Structured REDCap training in Tanzania demonstrated high user satisfaction and improved data quality, showing that targeted capacity-building can accelerate the adoption of secure data management systems in low-resource settings. These findings highlight the crucial role of training in bridging digital infrastructure gaps and enhancing sustainable biomedical research capacity across Africa.

## Materials and Methods

### Study Setting

This study was conducted within the Muhimbili Sickle Cell Program (MSCP), an initiative of Muhimbili University of Health and Allied Sciences (MUHAS). The MSCP addresses sickle cell disease through comprehensive research, patient care, training, and advocacy, collaborating with the Human Heredity and Health in Africa Bioinformatics Network (H3ABioNet) [19] and other international partners, including the Sickle Pan-African Research Consortium (SPARCo). These partnerships provide a substantial backdrop for deploying and assessing the utility of REDCap in enhancing clinical and research data management practices. MUHAS also serves as a regional training hub for health informatics and biomedical research, positioning it as an early adopter of REDCap in East Africa.

### REDCap Implementation

MUHAS secured a REDCap license from Vanderbilt University in 2011 and initiated phased implementation within the Muhimbili Sickle Cell Program (MSCP). The system was first piloted in 2014 on a test server operated on a non-traditional desktop, which enabled gradual staff training and orientation. The 2015 expansion increased familiarity with the platform, and by 2016, a dedicated institutional server was acquired, providing enhanced capacity for secure data management and research activities. The institutional server emphasizes robust data integrity, improved security protocols, and tailored user account management, thereby strengthening project administration. In 2017, REDCap expanded beyond the Muhimbili Sickle Cell Program to include the Sickle Pan-African Research Consortium (SPARCo) registry. By January 2023, the registry was deployed across 11 satellite sites in Tanzania (Figure 1) to support sickle cell research and patient follow-up. This scale-up demonstrates the scalability and adaptability of REDCap for multisite research coordination in resource-limited settings [15]. The expansion also provides a baseline for assessing REDCap’s adoption trajectory and its broader impact on institutional research capacity.

**Figure 1:**
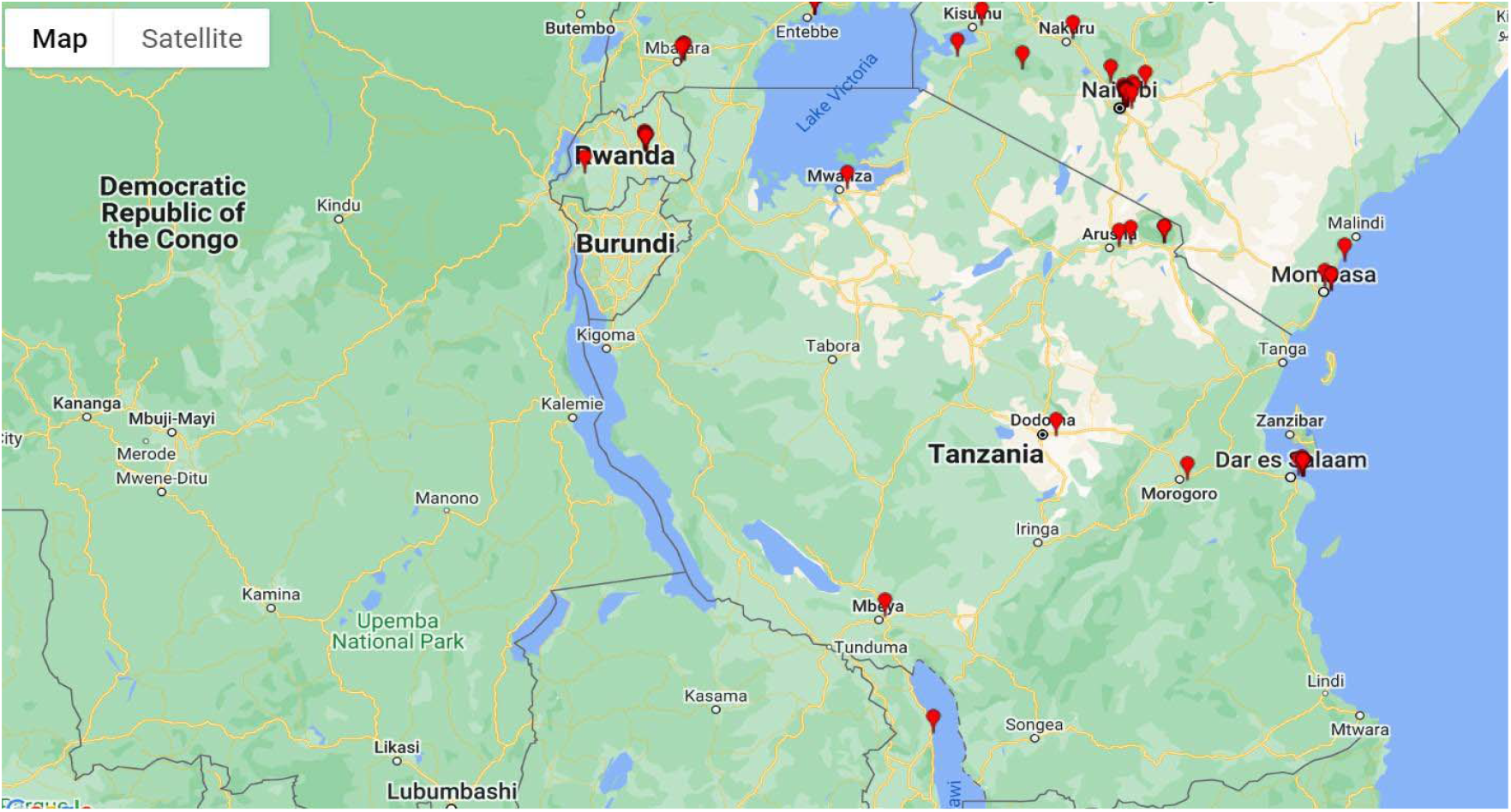
Geographic distribution of sites in Tanzania currently using REDCap as of January 2023

### User support and training

Ongoing technical support for REDCap users at MUHAS included creating and managing user accounts, troubleshooting, and ensuring data quality. To enhance user proficiency, the MSCP has developed a comprehensive training program that covers project setup, data entry, data management, and quality control. Training was delivered through structured workshops, which included practical sessions and hands-on exercises using a dedicated “training instance” of REDCap. This allowed trainees to practice without interfering with live projects, thus minimizing errors in production environments. Between 2016 and 2022, six REDCap training sessions were conducted with a total of 120 participants. These included academic and research staff, postgraduate and undergraduate students, collaborators from Tanzanian institutions, and regional participants from Ghana and Nigeria, reflecting both local and cross-border capacity-building efforts.

Training effectiveness was evaluated using a structured REDCap-based feedback survey comprising demographic items (gender, age, institution, cadre), baseline digital literacy (Microsoft Office, statistical packages, literature search), prior exposure to REDCap, and multiple 5-point Likert-scale items assessing satisfaction, perceived usefulness of training, and REDCap’s potential to improve research efficiency, institutional visibility, and career development.

### Data collection and measures

Institutional REDCap server audit logs, project metadata, user activity data, and training registration records were analyzed between 2016 and January 2023. The project-level measures included the number of projects created, their stage (development versus production), purpose, and use of survey instruments. Data capture was assessed by counting the number of instruments, fields, records, and submitted survey responses. Feature utilization was evaluated using double-entry verification, randomization modules, and mobile applications. User engagement was assessed based on the number of accounts created, active or inactive status, suspension due to prolonged inactivity and the number of events logged. The training data included participant demographics and survey responses collected during the workshops.

Survey data were exported directly from REDCap using the structured instrument described above, ensuring consistency and minimizing transcription errors.

### Definitions and analysis

Active users were defined as those performing substantive actions (e.g., project creation or data entry), whereas inactive users logged in without contributing any activity. Suspended users are locked because of prolonged inactivity. Projects in “production” were considered live, whereas those in “development” were still in testing. Descriptive statistics summarized adoption trends, training outcomes, and feature utilization, while temporal trends in project growth and user activity were visualized graphically.

### Ethics Statement

Ethical review was not required for this educational evaluation, as per the institutional guidelines. Participant consent was obtained for the survey responses.

## Results

### Project adoption

Between 2016 and 2023, 125 REDCap projects were created at MUHAS. Of these, 23 (18.4%) were initiated using the project template, 106 (84.8%) remained in development, and 19 (15.2%) were advanced to production status. Nearly half of the projects (63, 50.4%) incorporated surveys, whereas 62 (49.6%) did not. In terms of project purpose, 56 (44.8%) were research-focused, 56 (44.8%) supported operational functions, 7 (5.6%) addressed quality improvement, and 6 (4.8%) were categorized as other. Project initiation increased steadily, exhibiting an exponential growth trend over the seven years (Figure 2).

**Figure 2:**
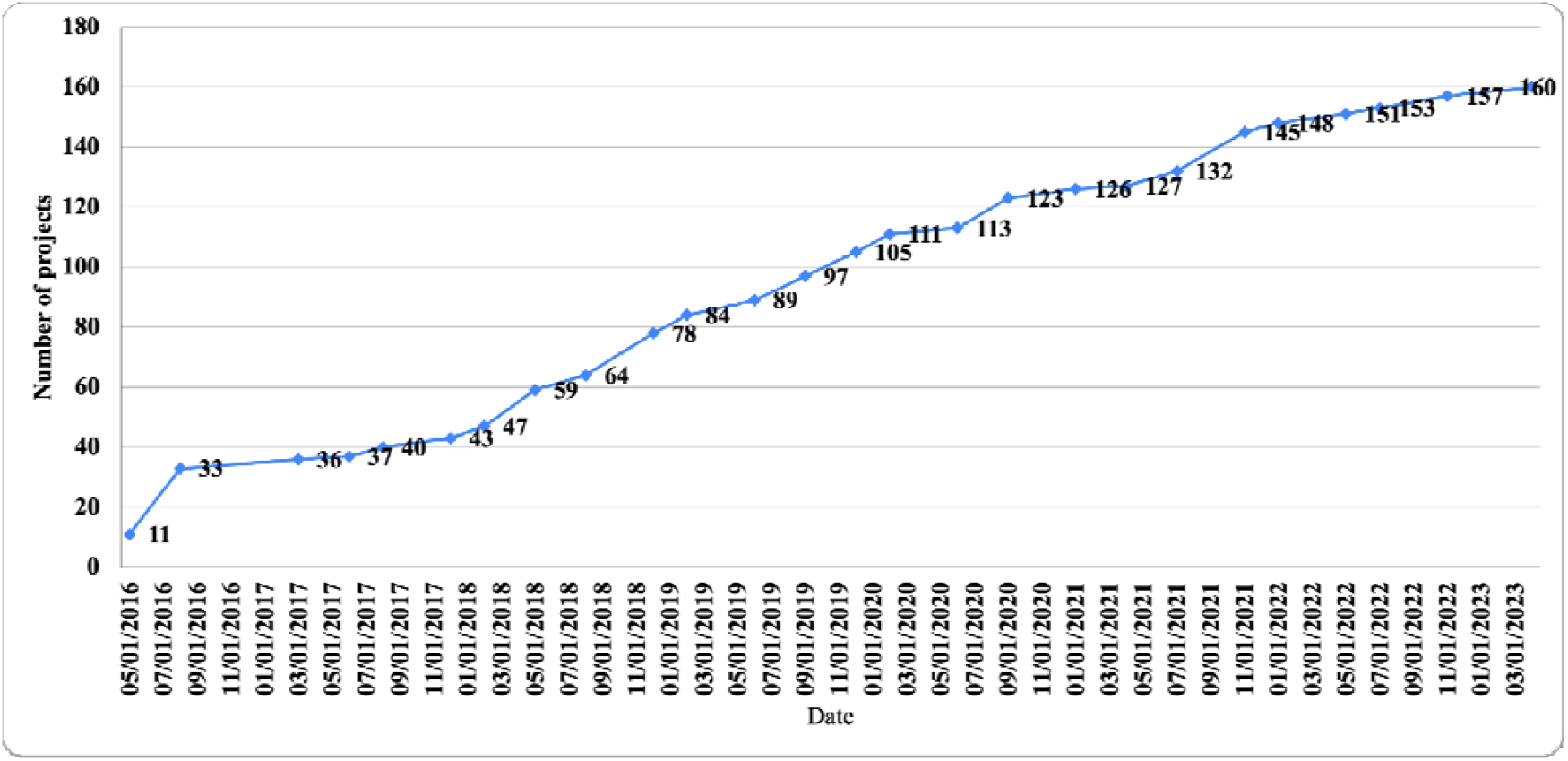
Annual trend in the number of projects created in REDCap between 2016 and 2023.

Across all projects, 601 data collection instruments were developed, comprising 16,040 unique fields and supporting 48,009 records (Table 1). A total of 4,197 survey responses were received, representing 491 unique participants among 119 survey invitations sent directly from REDCap. Of these, 45 (37.8%) generated responses, while 74 (62.2%) did not. Additional responses were collected through alternative distribution channels, including email and social media. The use of advanced features was limited: only two projects employed double data entry, and one project used the randomization module. No projects have implemented the Twilio telephony service.

**Table 1:** Attributes of projects created within REDCap in MUHAS, Tanzania, between 2016 and 2023.

| **Project attributes (n=125)** |  |
| --- | --- |
| Data collection instruments | 601 |
| Fields on all data collection instruments | 16,040 |
| Records | 48,009 |
| Survey responses | 4,197 |
| Survey participants (both invited from REDCap, email and others) | 491 |
| Survey invitations sent | 119 |
| Responded | 45 |
| Not responded | 74 |
| Double entry | 2 |
| Randomization | 1 |
| Twilio telephone service for the survey | 0 |

### System feature utilization

Mobile app functionality was used by 56 individuals across 32 projects, of which only 24 imported data via the app. By January 2023, the system had logged 378,413 discrete events, including 2,574 in the past seven days and 6,752 in the preceding 30 days.

### User engagement

By January 2023, 218 user accounts had been registered, of which 157 (72.0%) were active, 17 (7.8%) were inactive, and 44 (20.2%) were suspended for long-term inactivity (Table 2) (Supplementary Figure 1). Active users engaged in substantive activities, such as project creation and data entry, whereas inactive users primarily logged in without further activity. Overall, 6,752 login events were recorded between December 2022 and January 2023. Trends in logged events and active users demonstrated consistent growth (Figure 3). The system storage requirements reached 2.50 GB on the MySQL server and 4.10 GB on the web server.

**Table 2:** Project- and user-level characteristics of REDCap adoption as of January 2023.

|  |  |
| --- | --- |
| **Total logged events** | 378,413 |
| Past 7 days | 2,574 |
| Past 30 days | 6,752 |
| **User Statistics** |  |
| Total user accounts | 218 |
| Active users | 157 |
| Nonactive users (excluding suspended users) | 17 |
| Suspended users | 44 |
| Median # users per project (prod/analysis-cleanup) | 1 |
| Users (table-based authentication) | 217 |
| **Project Characteristics** |  |
| Projects not containing surveys | 1 |
| Projects containing surveys | 2 |
| Avg/median projects a user can access | 2.2/1 |

**Figure 3:**
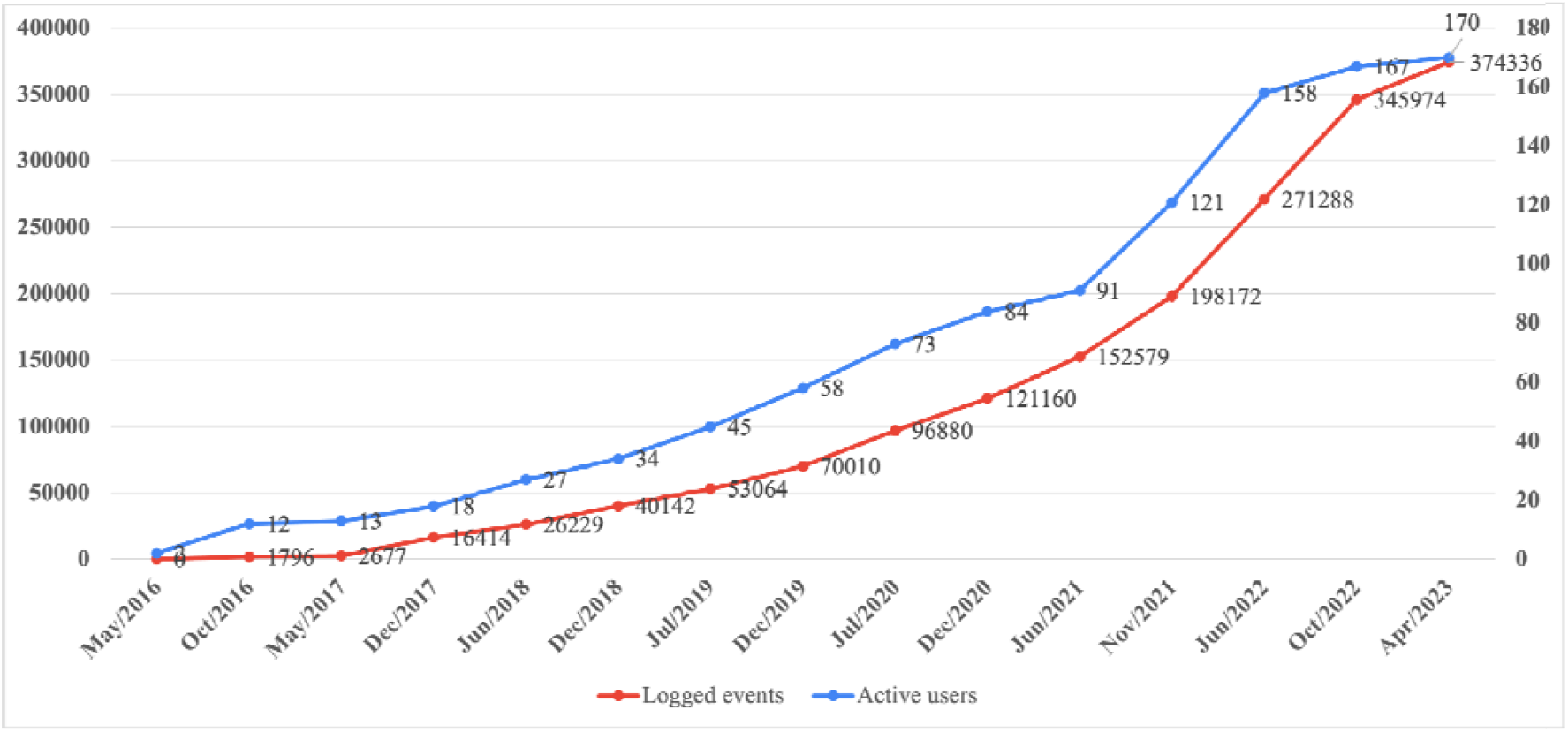
Trends in active user accounts and logged system events over time.

This growth was attributed to user activities and events (Table 2). The other events are shown in Supplementary Figure 1.

### Training outcomes

Between 2016 and 2022, six structured REDCap training sessions were conducted with 120 participants, of whom 110 completed the post-training evaluation. Most of the trainees were male (69, 62.7%), and the majority (58, 52.7%) were aged 26–32 years. Participants were drawn from the MUHAS (73, 66.4%), with additional representation from other Tanzanian institutions and partner sites in Ghana and Nigeria (Figure 4). By cadre, 46 (41.8%) were academic or research staff, 31 (28.2%) were postgraduate students, 13 (11.8%) were undergraduate students, and 20 (18.2%) were from other professional backgrounds.

**Figure 4:**
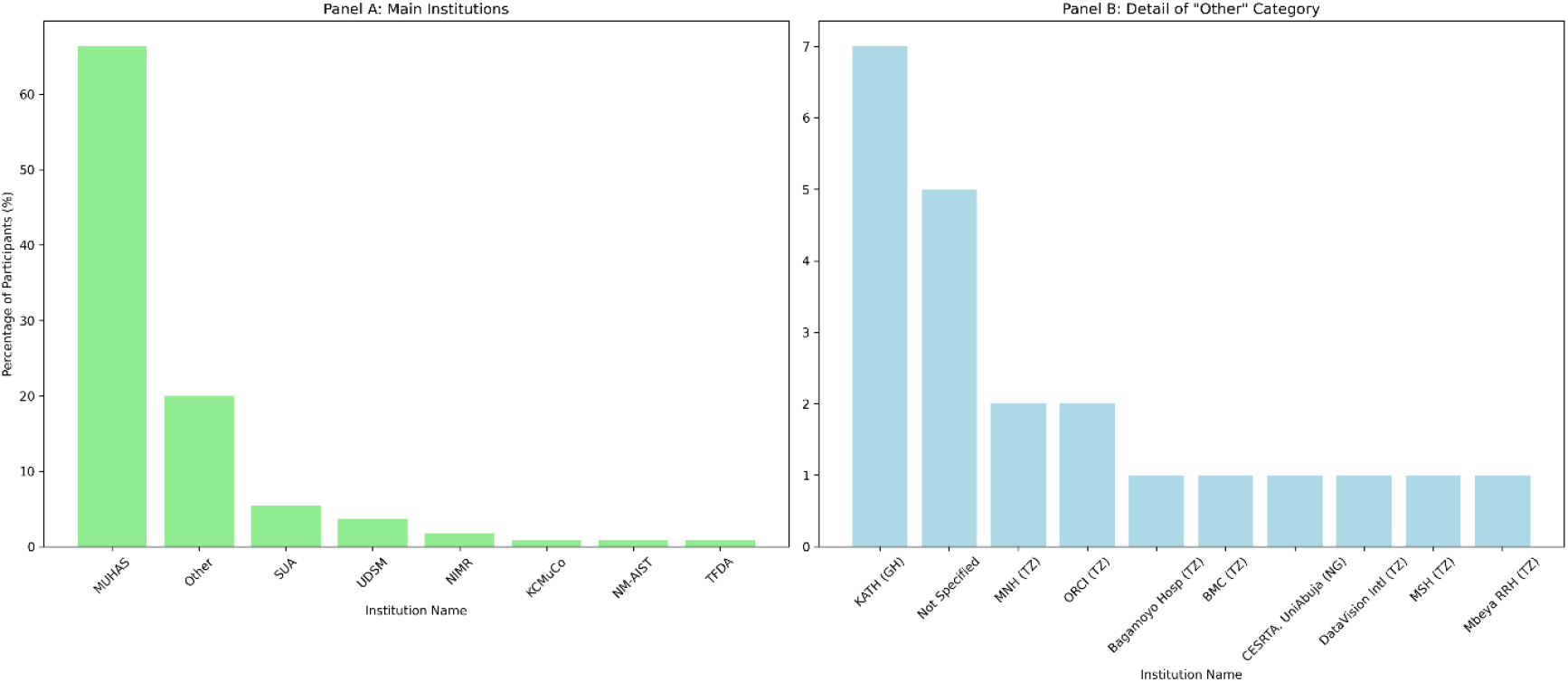
Institutional distribution of REDCap training participants, including Tanzanian sites (TZ) and regional partners in Ghana (GH) and Nigeria (NG). Panel A’s leading institutions are mostly from Tanzania, except for the categories expanded in Panel B, which include institutions from Tanzania, Ghana, and Nigeria. Key: MUHAS = Muhimbili University of Health and Allied Sciences, Tanzania; KCMuCo = Kilimanjaro Christian Medical University College, Tanzania; NIMR = National Institute for Medical Research, Tanzania; NM-AIST = Nelson Mandela African Institute of Science and Technology, Tanzania; SUA = Sokoine University of Agriculture, Tanzania; TFDA = Tanzania Food and Drug Authority; UDSM = University of Dar es Salaam, Tanzania; TZ = Tanzania; GH = Ghana; NG = Nigeria. Other Institutions: Bagamoyo Hosp, TZ = Bagamoyo District Hospital, Tanzania; Bugando MC, TZ = Bugando Medical Centre, Mwanza, Tanzania; CESRTA, UniAbuja, NG = Centre of Excellence for Sickle Cell Research and Training, University of Abuja, Nigeria; DataVision Intl, TZ = DataVision International Limited, Tanzania; KATH, GH = Komfo Anokye Teaching Hospital, Kumasi, Ghana; MSH, TZ = Management Sciences for Health, Tanzania; MNH, TZ = Muhimbili National Hospital, Dar Es Salaam, Tanzania; ORCI, TZ = Ocean Road Cancer Institute, Dar Es Salaam, Tanzania; Mbeya RRH, TZ = Mbeya Regional Referral Hospital, Mbeya, Tanzania.

Post-training evaluations consistently indicated high satisfaction across all REDCap use domains. Mean scores exceeded 4.4 on the 5-point Likert scale, with trainees strongly endorsing REDCap’s potential to enhance research effectiveness (mean 4.53, SD 0.65), increas productivity (mean 4.52, SD 0.69), and facilitate task completion (mean 4.50, SD 0.68). More than 85% of the participants agreed or strongly agreed with these statements (Figure 5), underscoring the acceptance of REDCap as a reliable and effective data management tool. Neutral or negative responses were rare, suggesting limited resistance to adoption. This pattern highlights both the immediate value of training and the perceived relevance of REDCap in strengthening institutional research capacity and improving study efficiency.

**Figure 5.**
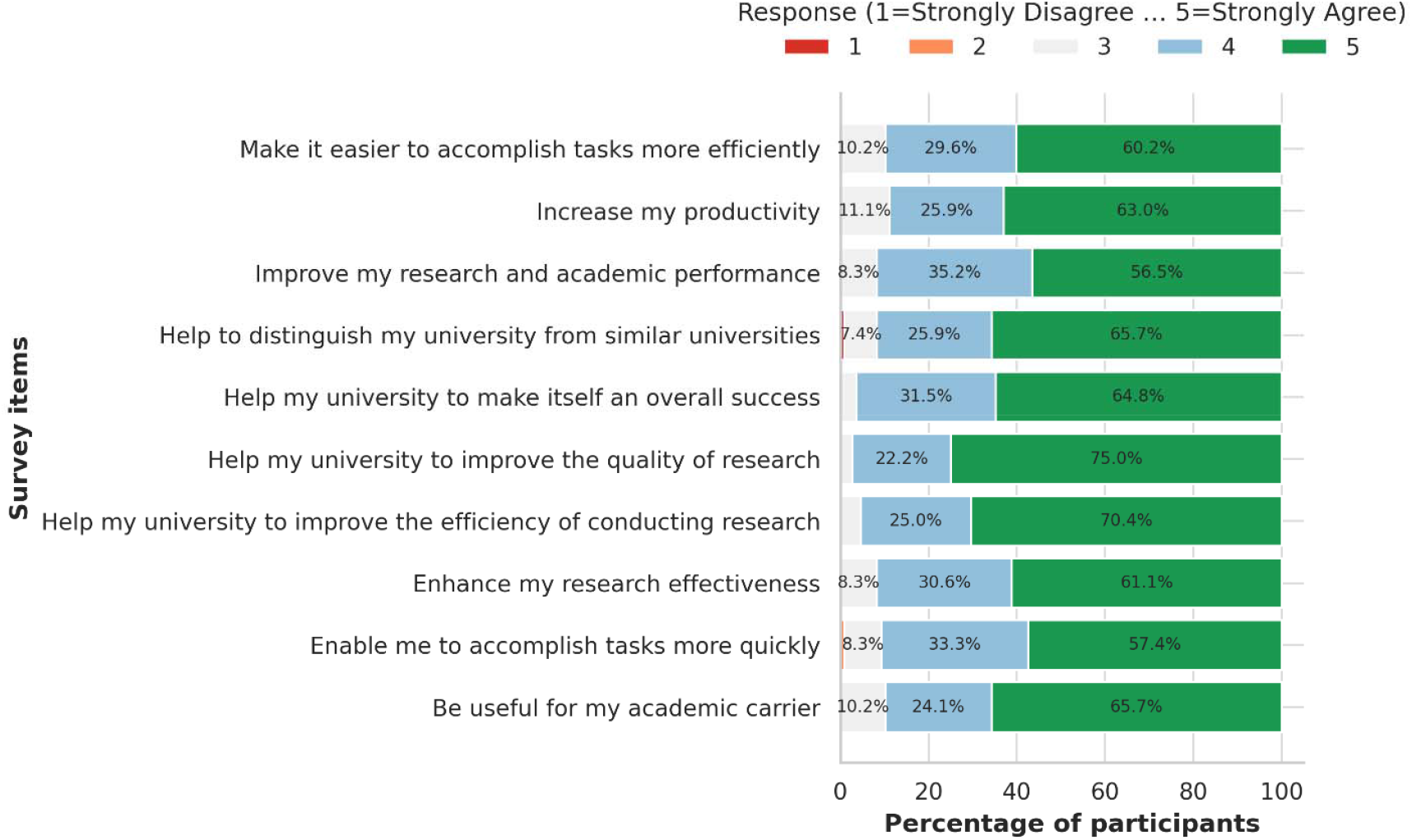
Diverging stacked bar chart of post-training evaluation responses on REDCap’s utility. Items are ordered by the mean score. Response options ranged from “Strongly disagree” (1) to “Strongly agree” (5).

## Discussion

This study charts the integration of REDCap into a Tanzanian institution’s research ecosystem, documenting its growth across 125 projects over seven years. Despite this expansion, survey engagement was limited, with only 45 of the 119 invited participants responding (37.8%). Such discrepancies echo the challenges seen in other low-resource settings, where participant follow-up is often inconsistent [9]. Future strategies should explore integrating mobile platforms and social media for survey distribution, as these tools have proven effective in enhancing response rates [8,16]. In parallel, attention to data privacy is essential, as apprehension over confidentiality may contribute to reluctance to respond. REDCap’s strong security framework [17] can mitigate such concerns; however, ongoing user training remains critical for ensuring both technical adoption and trust.

In Tanzania, researchers have traditionally relied on tools such as Excel, SPSS, KoboToolbox, Google Forms, and OpenClinica for data management [8,9,18]. Although these tools are familiar and accessible, they often lack advanced features for multisite collaboration, secure data access, and integration with clinical systems. In contrast, REDCap offers structured project workflows, audit trails, and customizable user permissions, making it particularly suited for clinical and academic research environments.

A pivotal advancement was the 2017 integration of REDCap into the Sickle Pan-African Research Consortium (SPARCo) registry, which linked 11 Tanzanian sites, including regional and district hospitals across Dar es Salaam, Pwani, Mwanza, and Zanzibar (Figure 1). This multisite expansion underscored REDCap’s adaptability in resource-limited environments and highlighted its role in harmonizing data collection for the longitudinal follow-up of sickle cell disease [20,21]. Compared with other multisite registries, such as SickleInAfrica [19] and regional pneumococcal studies [20], REDCap enables decentralized data capture while maintaining central oversight, providing project-specific workspaces that minimize interference across projects [5,21]. Its multisite accessibility and centralized project creation supported uniform quality control across networks, a key factor for research reliability in collaborative African initiatives.

Between 2016 and 2022, six REDCap training workshops were conducted, with 120 participants and 110 participants completing follow-up surveys. The demographic distribution—62.7% male, with the majority aged 26–32 years and representing academic staff, postgraduate, and undergraduate students—illustrates REDCap’s appeal to early- and mid-career professionals. Most participants were from MUHAS (66.4%), but their representation also extended to institutions in Ghana and Nigeria, fostering cross-border collaboration. This aligns with broader capacity-building efforts in African biomedical research [22–24]. Training evaluations revealed high satisfaction, with participants consistently highlighting the potential of REDCap to improve data quality, efficiency, and institutional visibility. Structured training has been proven essential for reducing errors, promoting standardization, and mitigating resistance to change, which are commonly encountered barriers to the adoption of new informatics tools [4,10,11].

The inclusion of participants from institutions across Tanzania, Ghana, and Nigeria illustrates the regional impact of training and its role in fostering cross-cultural exchange of data management practices. This diverse participation enriches the training experience and encourages collaborative research efforts across African institutions. By leveraging REDCap’s comprehensive data management tools, these training sessions have laid the groundwork for future research initiatives, promising to elevate research standards and data handling across networks and continents. Such training has profound implications, and standardized quality control procedures can potentially lead to improved research outcomes [5,7]. Ghana, Nigeria, and Tanzania collaborated with the SPARCo Project during the training period. This was followed by countries that adopted REDCap implementation in their own countries [19,23,25– 27].

Training evaluations revealed consistently high satisfaction, with mean scores exceeding 4.4 across all domains. The participants emphasized REDCap’s potential to improve data quality, research efficiency, and institutional visibility. These findings reinforce the importance of structured training not only for technical skill development but also for overcoming barriers frequently reported in new informatics implementations, including errors, lack of standardization, and resistance to change [10,11,13]. Such capacity-building efforts are indispensable for sustaining data-driven research and advancing the biomedical sciences across Africa.

Our results illustrate that structured training can accelerate adoption at the institutional level. However, scaling this impact requires sustained funding and investment in data science infrastructure across Africa [14,28]. Without parallel policies and funding commitments, isolated institutional initiatives cannot achieve long-term sustainability.

Beyond project adoption and training, REDCap’s interoperability features position it as a foundation for future data reuse and integration in Tanzania. The Shared Data Instrument Library (SDIL), curated by the REDCap Library of Official Content (REDLOC), enables the reuse of standardized forms and ensures harmonized data dictionaries across projects [29]. The introduction of “bottom-up” instrument sharing further enhances collaboration, allowing locally developed instruments to be disseminated after expedited review. These functions promote efficiency and facilitate Pan-African data harmonization in areas such as sickle cell research.

Interoperability with clinical systems provides an even broader frontier. REDCap’s integration with Fast Healthcare Interoperability Resources (FHIR) standards has established it as a leader in clinical data interoperability services [3]. Features such as Clinical Data Pull (CDP) enable the selective import of EHR data with adjudication for quality control. Simultaneously, Clinical Data Mart supports bulk extraction, which facilitates integration with major EHR platforms (e.g., Cerner and Epic) [5,7]. Additionally, REDCap2SDTM facilitates the transformation of project data into the Clinical Data Interchange Standards Consortium (CDISC) format, ensuring regulatory submission compatibility and enabling the reuse of clinical trial data [30]. Given the sensitivity of medical research data, REDCap’s built-in features, such as role-based access control, audit logs, and encryption, help safeguard data integrity while supporting secure sharing across projects [17]. These security measures are critical when balancing interoperability requirements with participant privacy, particularly in multisite and cross-border collaborations. Together, these features position REDCap as both a research data management tool and a sustainable repository for data reuse, with the potential to drive future data science initiatives in Tanzania.

The Tanzanian experience resonates with the international applications of REDCap in complex, multisite research. For example, the NHLBI-funded Sickle Cell Disease Implementation Consortium (SCDIC) registry used REDCap to manage standardized data collection for 2,400 patients across eight U.S. sites, demonstrating its scalability and ability to harmonize instruments across networks [31]. Similarly, the Australian Hemoglobinopathy Registry used REDCap to collect clinical and demographic data from 359 patients, demonstrating its adaptability to high-income settings with diverse patient populations [32]. The Globin Research Network for Data and Discovery (GRNDaD) registry collected data on 570 adults across 11 sites, further underscoring the role of REDCap in enabling large-scale, longitudinal, and collaborative research on sickle cell disease [33]. These global case studies reinforce REDCap’s versatility and highlight lessons applicable to Tanzania, particularly the importance of standardization, interoperability, and sustained investment in training.

### Limitations

Despite these successes, this study had several limitations. Infrastructure constraints, including intermittent Internet connectivity, hindered real-time data entry, while server hosting and hardware acquisition introduced ongoing costs. Resistance to change among staff members accustomed to legacy tools also challenged their adoption. Consistency in multisite data entry requires continued oversight, and user access management creates an additional administrative burden. Although REDCap provides strong security and access controls, balancing interoperability with privacy concerns remains an area that requires user training and education. Importantly, this evaluation did not capture patient- or staff-level perspectives on digital adoption, which may have influenced the scalability of the initiative. Finally, while REDCap use was associated with perceived gains in productivity and data quality, the absence of a baseline limited our ability to quantify improvements over time.

## Conclusion

Over the past seven years, REDCap adoption at MUHAS and across Tanzanian sites has transformed research data management, supporting 125 projects, multisite collaborations, and regional training initiatives. Its versatility, spanning secure survey capture, multisite accessibility, interoperability with clinical systems, and data reuse, positions it as a cornerstone of research infrastructure in Tanzania and beyond. Future development should focus on leveraging REDCap for data science, incorporating artificial intelligence and machine learning for advanced analytics, and designing more user-friendly interfaces to support non-technical users. Investment in training and infrastructure is essential to realizing REDCap’s full potential in advancing research quality, productivity, and impact across Africa.

## Data Availability

The data underlying this study are available without restriction, as they do not contain patient-identifying information. De-identified survey metadata can be obtained from the corresponding author upon reasonable request.

## Supplementary Material

Supplementary material is attached.

## Ethics Issues

Participants in the training provided feedback after providing their consent and were assured that no identifying information would be recorded.

## Author Contributions

All authors participated in writing and revising the manuscript.

## Supplementary Materials

## Supplementary Figure

**Supplementary Figure 1:**
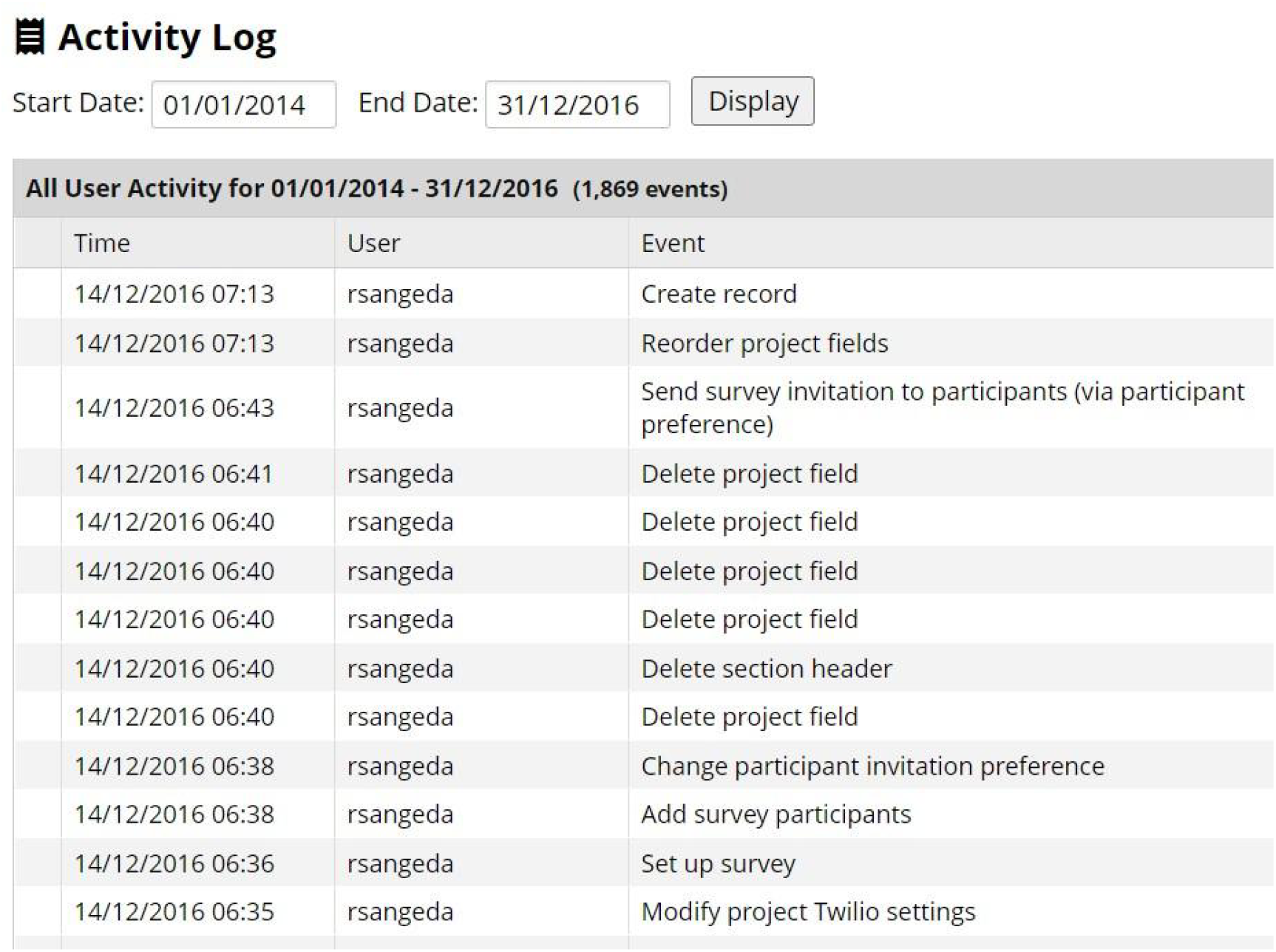
Early activities evidence of logged events in REDCap - truncated to show a few events from the REDCap server events log.

## References

Mayer MA, Furlong LI, Torre P, et al. Reuse of EHRs to Support Clinical Research in a Hospital of Reference. Stud Health Technol Inform. 2015;210:224–6.

Bastião Silva LA, Días C, van der Lei J, et al. Architecture to Summarize Patient-Level Data Across Borders and Countries. Stud Health Technol Inform. 2015;216:687–90.

Cheng AC, Duda SN, Taylor R, et al. REDCap on FHIR: Clinical Data Interoperability Services. J Biomed Inform. 2021;121:103871. doi: 10.1016/j.jbi.2021.103871

REDCap Consortium. REDCap project. https://www.project-redcap.org/software (accessed 30 September 2025)

Harris PA, Taylor R, Minor BL, et al. The REDCap consortium: Building an international community of software platform partners. J Biomed Inform. 2019;95:103208. doi: 10.1016/j.jbi.2019.103208

Garcia KKS, Abrahão AA. Research Development Using REDCap Software. Healthc Inform Res. 2021;27:341–9. doi: 10.4258/hir.2021.27.4.341

Harris PA, Delacqua G, Taylor R, et al. The REDCap Mobile Application: a data collection platform for research in regions or situations with internet scarcity. JAMIA Open. 2021;4. doi: 10.1093/jamiaopen/ooab078

Maduka O, Akpan G, Maleghemi S. Using Android and Open Data Kit Technology in Data Management for Research in Resource-Limited Settings in the Niger Delta Region of Nigeria: Cross-Sectional Household Survey. JMIR Mhealth Uhealth. 2017;5:e171. doi: 10.2196/mhealth.7827

King C, Hall J, Banda M, et al. Electronic data capture in a rural African setting: evaluating experiences with different systems in Malawi. Glob Health Action. 2014;7. doi: 10.3402/gha.v7.25878

Maré IA, Kramer B, Hazelhurst S, et al. Electronic Data Capture System (REDCap) for Health Care Research and Training in a Resource-Constrained Environment: Technology Adoption Case Study. JMIR Med Inform. 2022;10:e33402. doi: 10.2196/33402

Klipin M, Mare I, Hazelhurst S, et al. The Process of Installing REDCap, a Web Based Database Supporting Biomedical Research. Appl Clin Inform. 2014;05:916–29. doi: 10.4338/ACI-2014-06-CR-0054

Sangeda RZ, Mwakilili AD, Masamu U, et al. A Baseline Evaluation of Bioinformatics Capacity in Tanzania Reveals Areas for Training. Front Educ (Lausanne). 2021;6:25. doi: 10.3389/feduc.2021.665313

Mwita LA, Mawalla WF, Mtiiye FR, et al. Infrastructure for bioinformatics applications in Tanzania: Lessons from the Sickle Cell Programme. PLoS Comput Biol. 2023;19:e1010848. doi: 10.1371/journal.pcbi.1010848

Kayalioglu H, Pateña J, Sangeda RZ, et al. The importance of funding and investment to strengthen data science in Africa. Communications medicine. 2025;5:293. doi: 10.1038/s43856-025-01001-7

Masamu U, Sangeda RZ, Mgaya J, et al. Improved Biorepository to Support Sickle Cell Disease Genomics and Clinical Research: A Practical Approach to Link Patient Data and Biospecimens from Muhimbili Sickle Cell Program, Tanzania. Biopreserv Biobank. 2024;22:248–56. doi: 10.1089/bio.2023.0060

Romeiser JL, Cavalcante J, Richman DC, et al. Comparing Email, SMS, and Concurrent Mixed Modes Approaches to Capture Quality of Recovery in the Perioperative Period: Retrospective Longitudinal Cohort Study. JMIR Form Res. 2021;5:e25209. doi: 10.2196/25209

IHR REDCap. REDCap Security overview. http://kpco-ihr.org/redcap/citing.html#twilio (accessed 30 September 2025)

Löbe M, Meineke F, Winter A. Scenarios for Using OpenClinica in Academic Clinical Trials. Stud Health Technol Inform. 2019;258:211–5.

Makani J, Sangeda RZ, Nnodu O, et al. SickleInAfrica. Lancet Haematol. 2020;7:e98–9. doi: 10.1016/S2352-3026(20)30006-5

Brown BJ, Madu A, Sangeda RZ, et al. Utilization of Pneumococcal Vaccine and Penicillin Prophylaxis in Sickle Cell Disease in Three African Countries: Assessment among Healthcare Providers in SickleInAfrica. Hemoglobin. 2021;1–8. doi: 10.1080/03630269.2021.1954943

Harris PA, Taylor R, Thielke R, et al. Research electronic data capture (REDCap)--a metadata-driven methodology and workflow process for providing translational research informatics support. J Biomed Inform. 2009;42:377–81. doi: 10.1016/j.jbi.2008.08.010

Mulder NJ, Adebiyi E, Alami R, et al. H3ABioNet, a sustainable pan-African bioinformatics network for human heredity and health in Africa. Genome Res. 2016;26:271–7. doi: 10.1101/gr.196295.115

Adekile A, Anie KA, Hamda C Ben, et al. The Sickle Cell Disease Ontology: enabling universal sickle cell-based knowledge representation. Database. 2019;2019:1–12. doi: 10.1093/database/baz118

Munung NS, Nembaware V, de Vries J, et al. Establishing a Multi-Country Sickle Cell Disease Registry in Africa: Ethical Considerations. Front Genet. 2019;10:1–8. doi: 10.3389/fgene.2019.00943

Nnodu O, Madu A, Chianumba R, et al. Establishing a database for sickle cell disease patient mapping and survival tracking: The sickle pan-african research consortium Nigeria example. Front Genet. 2022;13. doi: 10.3389/fgene.2022.1041462

Paintsil V, Amuzu EX, Nyanor I, et al. Establishing a Sickle Cell Disease Registry in Africa: Experience From the Sickle Pan-African Research Consortium, Kumasi-Ghana. Front Genet. 2022;13:1–10. doi: 10.3389/fgene.2022.802355

Kandonga D, Sangeda RZ, Masamu U, et al. Development of the sickle Pan-African research consortium registry in Tanzania: opportunity to harness data science for sickle cell disease. Frontiers in Hematology. 2023;2:1–8. doi: 10.3389/frhem.2023.1040720

Sangeda RZ, Nkya S, Masamu U, et al. Funding Trends and Gender Disparities in Tanzanian Biomedical Research: Insights From Global Health Funding Databases. Cureus. Published Online First: 2025. doi: 10.7759/cureus.94912

Obeid JS, McGraw CA, Minor BL, et al. Procurement of shared data instruments for Research Electronic Data Capture (REDCap). J Biomed Inform. 2013;46:259–65. doi: 10.1016/j.jbi.2012.10.006

Yamamoto K, Ota K, Akiya I, et al. A pragmatic method for transforming clinical research data from the research electronic data capture ‘REDCap’ to Clinical Data Interchange Standards Consortium (CDISC) Study Data Tabulation Model (SDTM): Development and evaluation of REDCap2SDTM. J Biomed Inform. 2017;70:65–76. doi: 10.1016/j.jbi.2017.05.003

Glassberg JA, Linton EA, Burson K, et al. Publication of data collection forms from NHLBI funded sickle cell disease implementation consortium (SCDIC) registry. Orphanet J Rare Dis. 2020;15:178. doi: 10.1186/s13023-020-01457-x

Nelson A, Ho PJ, Haysom H, et al. Sickle cell disease in Australia: a snapshot from the Australian Haemoglobinopathy Registry. Intern Med J. 2024;54:764–72. doi: 10.1111/imj.16297

Boye-Doe A, Brown E, Puri-Sharma C, et al. The Grndad Registry: Contemporary Natural History Data and an Analysis of Real-World Patterns of Use and Limitations of Disease Modifying Therapy in Adults with SCD. Blood. 2020;136:34–6. doi: 10.1182/blood-2020-138895

